# Fostering inclusion in clinical and research procedures in children with neurodevelopmental disorders through augmentative and alternative communication: a narrative review and a proposal for visual aids application in brain stimulation

**DOI:** 10.1101/2024.10.31.24316252

**Authors:** Sara Passarini, Fabio Quarin, Giulia Lazzaro, Floriana Costanzo, Andrea Battisti, Giovanni Valeri, Silvia Guerrera, Laura Casula, Deny Menghini, Sabine Pirchio, Stefano Vicari, Elisa Fucà

## Abstract

**Background and aims:** Children with neurodevelopmental disorders (NDDs) such as Autism Spectrum Disorder (ASD) and Intellectual Disability (ID) may exhibit medical and neurocognitive comorbidities requiring periodic medical check-ups. In addition, children with NDDs often experience language difficulties that hamper an effective communication in the healthcare setting, with potential deleterious effects on their compliance. In turn, these challenges may affect accessing healthcare resources and receiving adequate short- and long-term assistance for their primary symptomatology and any related comorbidities. Communication impairments may also pose barriers for involving children with NDDs in clinical trials and research procedures, such as those based on the use of Non-Invasive Brain Stimulation (NIBS). NIBS has shown various benefits across different conditions and researchers are increasingly implementing its application, also involving children with ASD and other NDDs.

Augmentative and Alternative Communication (AAC) may be a valuable strategy to sustain the participations of children with NDDs in research procedures and trials, promoting ethical principles of equitable participation and inclusion while adopting a patient-centered approach.

This systematically-conducted review aimed to explore (i) the use of ACC to sustain compliance during research procedures, specifically focusing on neuromodulation procedure involving children with NDDs; (ii) the employment of AAC tools to sustain the compliance of children with NDDs in clinical settings.

**Methods:** Articles were searched from PubMed database. All found articles were screened by title and abstract, followed by full text if they met the inclusion criteria.

**Results:** The first search yielded no studies investigating the use of AAC among children with NDDs to support comprehension and implementation of brain stimulation procedures. From the second launched search, twelve studies were reviewed. Almost the majority of eligible papers involved children with ASD (67%), including a small percentage of children with ID (8%) or with mixed etiology NNDs (25%). Of note, most of the included papers (8 out of 12) focused on the AAC use in the dental care setting.

**Conclusion:** Despite the methodological variability and the paucity of studies exploring the potential of AAC in fostering compliance with clinical and research procedures, this review highlights AAC’s potential for children with NDDs in decreasing stress and anxiety levels while boosting their adherence to healthcare practices. Starting from the current literature evidence, an AAC protocol for boosting compliance with NIBS in children with NDDs and language impairments is proposed.

**Implications:** Promoting the use ACC in clinical and research settings could be a beneficial approach to enhance clinical practice and evidence-based literature. By developing an AAC protocol for NIBS, our proposal aims to fill these gaps. Ensuring compliance with research procedures is crucial for delivering gold standard care, adhering to ethical statements while advancing scientific knowledge.

## 1 Introduction

NDDs are a group of heterogeneous conditions with onset during the early developmental and global functioning impairments (American Psychiatric Association, 2022). NDDs include Intellectual Disability (ID), Communication Disorders, Autism Spectrum Disorder (ASD), Attention-Deficit/Hyperactivity Disorder, Neurodevelopmental Motor Disorders, and Specific Learning Disorders (American Psychiatric Association, 2022).

Behavioral, cognitive, and psychological interventions, particularly those targeting communication and social skills, are widely use to address impairments of children with NDDs (Gómez et al., 2021; Hyman et al., 2020; Peterson et al., 2024). In contrast, pharmacological treatments are primarily focused on physical and mental health comorbidities (Hyman et al., 2020; Totsika et al., 2022; Van Vyve et al., 2023). Therefore, there is pressing need for new intervention options.

In recent decades, Non-Invasive Brain Stimulation (NIBS) techniques are attracting growing interest for the possibility to tackle at circuits’ level the pathophysiology of the different diseases (D’Urso et al., 2020). Among NIBS, Transcranial Direct Current Stimulation (tDCS) has emerged as a novel, cost-effective and research-based therapeutic technique for Neurodevelopmental Disorders (NDDs) (Faralli et al., 2024; Fertonani et al., 2015; Finisguerra et al., 2019; Palm et al., 2016; Prasad et al., 2024), with minor adverse events (Buchanan et al., 2021). For example, available research showed improvements in language skills, working memory, severity symptomatology, theory of mind, withdrawal, emotion regulation and challenging behaviors in children with ASD after tDCS sessions (D’Urso et al., 2015; Oberman et al., 2024). For individuals with Down Syndrome (DS), tDCS seems to provide enhancements in comorbid psychopathology (e.g. depressive and catatonic symptomatology), cognitive and motor abilities (e.g. planning and velocity) (Faralli et al., 2024; Ramezani Golafzani, N., Karami, A., & Rostami, R., 2021). Additionally, positive effects have been reported also for children with dyslexia (Cancer & Antonietti, 2018) and with Attention-Deficit/Hyperactivity Disorder, by enhancing attention and working memory skills (D’Urso et al., 2020). Given the benefits of NIBS for NDDs, it is crucial to implement these techniques (Prasad et al., 2024).

However, the existence of an *underrepresentation* bias excluding from clinical trials specific subgroups of individuals with NDDs, such us children with ID or ASD has been pointed out (Feldman et al., 2014; Russell et al., 2019). An estimated 90% exclusion rate of children with NDDs exists in child development research, despite the potential for their participation with adequate support tools, such as visual aids (Feldman et al., 2013, 2014). Additionally, the majority of research on ASD involved children with preserved language and cognitive abilities, excluding individuals with ASD and low cognitive functioning due to their challenging and noncompliant behaviors with trial procedures (Jack & A. Pelphrey, 2017). Brain stimulation research also tends to include children with ASD and good cognitive skills; on the other hand, available studies often do not provide data on participants’ cognitive functioning (Luckhardt et al., 2021). Similarly, researchers may be incline to exclude people exhibiting specific conditions, such as DS, due to their motor limitations and impulsivity that may make it difficult to comply with study demands (Baumer et al., 2022).

In accordance with the Helsinki ethical statement for researchers (2008), equal opportunities and access must be guaranteed (Muñoz & Borbón, 2023). Clinical trials represent the most robust studies for advancing scientific knowledge regarding the effectiveness of new drugs or interventions. Adopting a restrictive research approach has important ethical, social, and scientific consequences (Feldman et al., 2014). This issue is particularly relevant for children with NDDs as they seem to exhibit high rates of mental and physical comorbidities (Al-Beltagi, 2021; Antonarakis et al., 2020; Kidd et al., 2014; Sigafoos et al., 2003), that should be addressed with all potential evidence-based interventions.

Of note, one of the barriers to inclusion in research is communication difficulties, which often affects individuals with NDDs (Jagoe et al., 2021). NDDs show complex phenotypes in which severe language impairments are often observed (Reindal et al., 2023; Tager-Flusberg, 2003). For instance, individuals with ASD, beyond the core symptoms of restrictive and repetitive behaviors and social communication impairments (American Psychiatric Association, 2022), may display language difficulties, leading to potential delays in its onset, regression of previously acquired skills, or cessation of further language development (e.g. language plateau). Receptive, expressive and pragmatic difficulties are noted at varying levels of severity (Schaeffer et al., 2023). Children with ID and genetically defined ID, such as DS and Fragile X syndrome, frequently display atypical speech development with expressive, receptive and pragmatic anomalies (Tūbele & Landrāte, 2021).

Since language is a complex system allowing people to join actively their daily life and to communicate their own needs, thoughts and feelings, impaired language skills may hesitate in emotional dysregulation in children with NDDs (Burnley et al., 2023). The fear of equipment and language impairment may hamper compliance with clinical and research demands leading to behavioral problems and noncompliance with clinical and research procedures (Cuvo et al., 2010). For instance, children with DS often struggle with behavioral dysregulation, attention, cognition, anxiety, and previous stressing experiences in clinical settings, all of which contribute to difficulties in tolerating the application of clinical and research instruments like polysomnography sensors (Collaro et al., 2023; Lanzlinger et al., 2023). Communication and behavioral difficulties may lead to the exclusion of individuals with NDDs from clinical trials, generating external validity bias and nourishing social disparities with ethical implications (Feldman et al., 2014; Russell et al., 2019). Restraint, sedation, or anesthesia could face noncompliance and challenging behaviors exhibited by children with NDDs in clinical setting, but with the risk of altering test results and amplifying their frustration (Paasch et al., 2012). Behavioral interventions (e.g. non-contingent reinforcement, differential reinforcements of other behavior, and differential reinforcement of alternative behavior) (Carter et al., 2019; Dufour & Lanovaz, 2020; Lillie et al., 2021; Piazza et al., 2015; Szalwinski et al., 2019; Tucker et al., 1998) may represent suitable alternatives.

Beyond these strategies, the communication barrier in clinical and research procedures could be overcome by using all modes of communication, including gestures and visual aids (Jagoe et al., 2021). Hence, Alternative and Augmentative Communication (AAC) may represent a valuable opportunity to enhance compliance of children with NDDs with clinical and research procedures.

### 1.1 Strategies to improve communication barriers: focus on Alternative and Augmentative Communication

AAC aims to improve communication by enhancing functional communication skills, language, social competence, and natural speech in individuals of all ages, regardless of the presence of a clinical condition (Crowe et al., 2022; Martínez-Santiago et al., 2018). AAC deals with two different meanings: augmentative, which applies to individuals with existing speech but limited intelligibility or expressive capabilities, and alternative to verbal language (Beukelman & Light, 2020; Brownlee & Palovcak, 2007). AAC encompasses various tools and strategies from sign language to external devices, such as alphabet boards, picture exchange communication systems (PECS), pictograms, and speech-generating device (International Society for Augmentative and Alternative Communication, 2020).

Direct modelling in real life situations is the most effective method to teach how to deal with AAC. When teaching the use of AAC, healthcare providers have to accompany their language with touching the symbol corresponding to the spoken word or using gestures to compose messages. Depending on the chosen AAC system, the healthcare provider may only touch or exchange pictograms with the child (e.g. PECS). Additionally, caregivers should join the introduction to AAC to familiarize with these tools with the aim of implementing communication opportunities in all life contexts (Beukelman & Light, 2020).

AAC boosts contextual language understanding, creating and arising communicative opportunities, and reducing interactions’ misunderstandings. Consequentially, AAC reduce communication barriers in various settings, included the clinical one (Blackstone & Pressman, 2016). Moreover, AAC has the potential to increase motivation and decrease challenging behaviors (Beukelman & Light, 2020; Light et al., 2019; Walker & Snell, 2013).

Due to ongoing positive advancements in the application of tDCS, it is crucial to consider all available strategies for enhancing its implementation. Since tDCS is a relatively new therapeutic technique, AAC schedules may be beneficial in helping children, parents, and staff become familiar with brain stimulation procedures and equipment.

Thus, aiming at implementing brain stimulation techniques while adhering to ethical principles of inclusivity for all potential beneficiaries and ensuring fully participation, the present study explored the effectiveness of AAC for supporting the application of brain stimulation procedures. Therefore, we launched an initial search exploring the use of AAC for supporting compliance during NIBS sessions. Given the lack of results, we conducted a second search focusing on AAC tools and compliance of children with NDDs within the healthcare context. Finally, we developed a set of visual aids to make brain stimulation easier to implement in children with NDDs and language impairment.

## 2 AAC and medical compliance in children with NDDs

### 2.1 Search questions

The present narrative review has explored the following aims:

1. Focusing on AAC inputs to sustain compliance during brain stimulation procedures in children with NDDs;
2. Focusing on AAC inputs to sustain medical compliance in children with NDDs.

## 3 Method

### 3.1 Search Procedure

An extensive literature search has been carried out for the present review using the database “PubMed” without starting time restrictions, until 6 January 2024. For the first search aim the following expressions were used: (“augmentative and alternative communication” OR “visual schedule” OR “picture schedules” OR “pictures”) AND (“intellectual disability” OR “autism” OR “communicative disability”) AND (“neuromodulation” OR “brain stimulation” OR “tDCS”). For the second aim, the first two algorithms were used adding the following term combination (“health care” OR “compliance” OR “medical setting”).

The first, second, and third authors conducted the literature search independently, screened the titles and abstracts of potentially eligible studies, examined the full texts, and extracted descriptive data, collaborating whenever the inclusion or exclusion of one study was doubtful.

### 3.2 Search selection

The articles obtained were selected according to different inclusion criteria, as follows:

a. articles written in English;
b. articles including children and adolescents with NDDs;

No time restriction was applied. Reviews of the literature were excluded.

## 4 Results

### 4.1 Research question 1

#### 4.1.1 Study selection

The first algorithm revealed 97 results. All found articles were screened for title and abstract; none of them focused on AAC, compliance and research procedures in general or specifically to brain stimulation in children with NDDs. The search did not find studies focusing on compliance with research procedures.

### 4.2 Research question 2

#### 4.2.1 Study Selection

The second algorithm revealed 497 results. Studies not in English language were excluded (n=10). Of the 487, 245 were excluded for not meeting the inclusion criteria of age population. Of the 242 remain studies, 18 reviews were excluded. 224 articles were screened for title and abstract; 211 were excluded. A total of 13 studies met inclusion criteria: each one of them was focused on pediatric population with NDDs and was written in English. Of the 13 studies, articles without pediatric clinical population were excluded (n=1). A total of 12 studies was revised (Figure 1).

**Figure 1.**
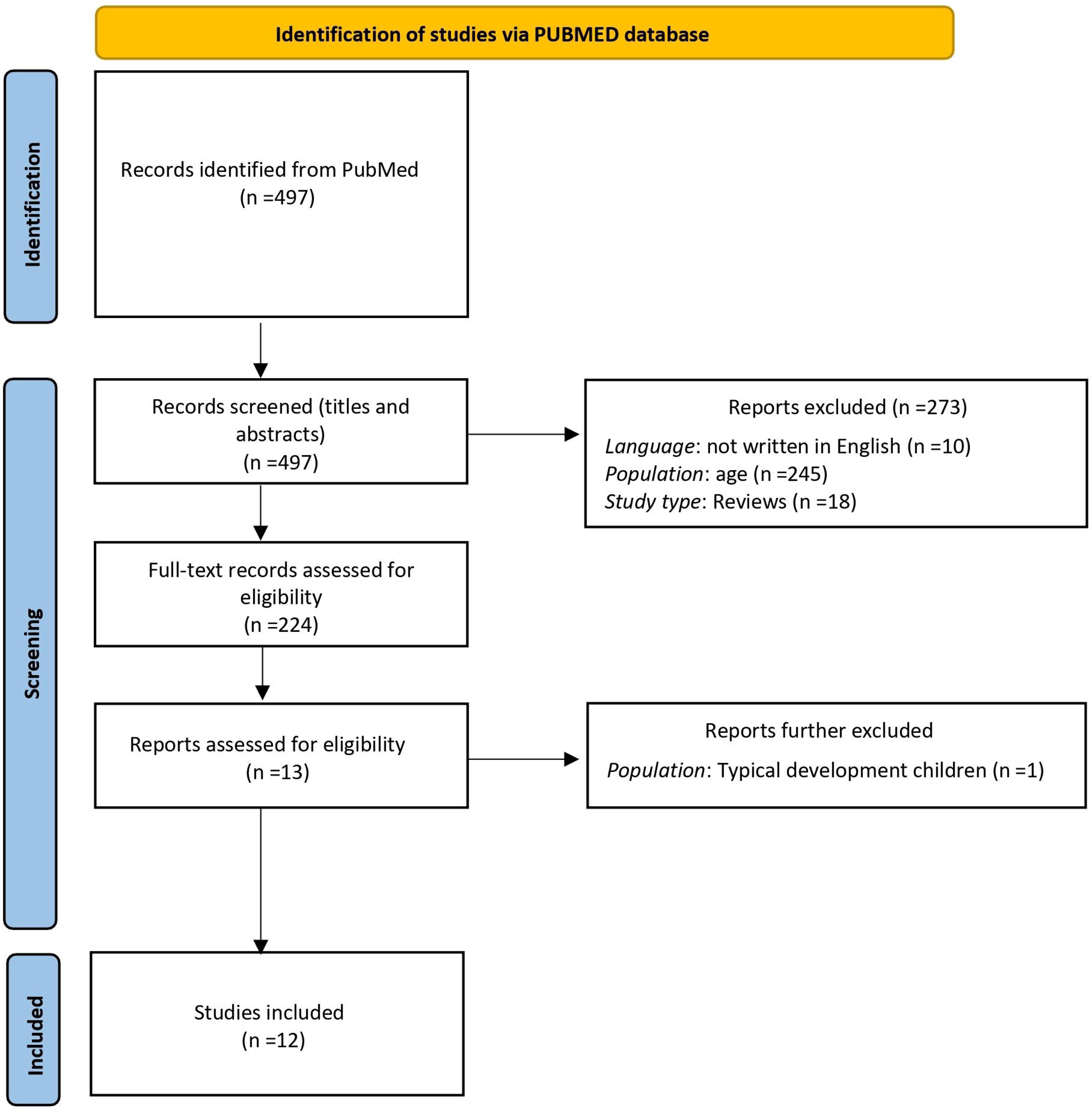
Search flow diagram of the second launched algorithm.

#### 4.2.2 Summary of Study Characteristics

The 67% of the studies (N=8) included in this narrative review were conducted in the dentistry context. In addition, the 67% (N=8) involved youth with ASD (age range 4-19 years), 8% focused on ID (N=1) and 25% on mixed NDDs (N=3). The study involving children with ID and one study including mixed NDDs do not specify population age.

#### 4.2.3 AAC and dental healthcare

One of the first studies conducted and included in this review by Isong and collaborators (Isong et al., 2014) used high tech AAC (video) to increase compliance and reduce fear during dental visits in 80 youth with ASD (age range 7-17 years; 81% males and 19% females). Children were assigned to different conditions: video peer modeling only (i.e. video of a child undergoing a preventive dental care visit from the arrive at the hospital to the oral care), video goggles only, video peer modeling plus video goggles, and control group. The Venham Anxiety and Behavior scale (Venham et al., 1980) highlighted decreased anxiety levels and more adaptive behaviors for video peer modeling plus video goggles condition (0.8 points); a greater reduction in anxiety was observed in children who saw the video peer more than once.

Grewal and collaborators (Grewal et al., 2015) evaluated the compliance for dental procedures in 60 children with mixed etiology NDDs, half part used low tech AAC while the other high-tech AAC tools. In addition, parents attended psychoeducational training to enhance children’s oral compliance. Both AAC sets described oral procedures (e.g. X-ray), rewards, and dental self-care activities (e.g. tooth brushing and mouthwash). Authors highlighted the efficacy of AAC boosted with parent training for all groups in statistically improving compliant behaviors and positive attitudes toward all dental exams, measured by the Frankl Behavior Rating Scale (S Frankl, F Shiere, H Fogels, 1962) and the Houpt behavior rating scale (Houpt et al., 1985).

Subsequently, Mah and Tsang (Mah & Tsang, 2016) used visual schedules to explain dental procedures to a group of 14 males with ASD (aged 4-8 years) who were non-compliant in past oral examinations. Half of the sample used AAC tools while the remaining children the tell-show-do method. Results underlined significant correlations between behavioral distress (e.g. screaming, crying, and escaping), assessed with the Child-Adult Medical Procedure Interaction Scale-Short Form (Blount et al., 2001), and the number of completed steps and the time required to end each procedure. Less distress levels were associated with a higher number of dental exams succeeded. Further, the number of steps completed increased with repeated weekly appointments for both groups, but children with visual cues ended oral medications with less amount of time.

Zink and colleagues (Zink et al., 2016) showed improvements in oral checkups using tailored pictograms in 26 children with ASD (mean age 10 ± 3.3 years; 22 males and 4 females). Of them, 13 children had no prior experience with dental treatments (G1), while 12 had previous experience (G2). G1 required significantly less time to perform specific medical steps using PECS. One subsequent study by Zink and collaborators (Zink et al., 2018) compared high tech AAC (e.g., app with images linked to audio descriptions) and low tech AAC (PECS) in enhancing communication during dental examinations involving 40 youth with ASD (aged 9 to 15 years; 38 males and 2 females), equally divided into 2 groups. Results indicated statistically significant differences in the number of attempts and the number of appointments for dental exams, with the app condition showing more improvements. Authors also suggested that a decrease in the number of appointments could potentially reduce reliance on general anesthesia for outpatient procedures.

These findings were confirmed by Lefer and colleagues (Lefer et al., 2019), who developed visual schedules of 6 dental pictograms for 52 children with ASD (mean age 10.2 years; 45 males and 7 females), combining and implementing the AAC tool with a psychoeducational program on oral health. Children underwent 5 dental care examinations (T0, T1, T2, T3, and T4) over 8-months in which their compliance with dental steps and their anxiety levels by using the Frankl Behavior Rating Scale (S Frankl, F Shiere, H Fogels, 1962) were monitored. Results highlighted improvements in tolerance towards dental examination: the estimated percentage ranging from 2 to 17% of children did not perform the first four steps at T0 decreased to 0% by the end of the search. The percentage of children achieving the initial 4 steps increased over time from a range of 37-79% (T0) to 75-86.5% (T4). The percentage of individuals mastering the entire exam procedure increased from 25% at T0 to 65.4% at T4. Additionally, the lack of perceived anxiety increased to 36.5% at T1, 51.9% at T2 and T3, and 59.6% at T4. The Global scores for compliance and anxiety levels showed significant differences between T0 and T1, as well as between T0 and T4.

Naidoo and colleagues (Naidoo & Singh, 2020) selected the most representative and child-friendly dental terms to create visual boards for clearly describing oral checkups to children with ASD (aged 7-14 years). In a survey with close and open-ended questions, the 80% of dental workers underlined the utility of AAC in explaining therapeutic plans to children with ASD and the 60% in boosting children’s compliance.

Myhren and collaborators (Myhren et al., 2023) investigated the efficacy of AAC tools (pictograms, photos or video) in improving oral checkup reducing the use of general anesthesia or sedation. 17 children with ASD (aged 6-13 years; 13 males and 4 females), 12 with partially or fully adequate language development and 15 with several sensory anomalies (e.g. sound, smell, and taste), underwent a “habituation program” for gradually introducing children to various dental examinations (e.g. X-ray). Results showed that the 82% of children completed all dental examination’s steps and the 59% underwent X-ray with good compliance, evaluated by the Frankl Behavior Rating Scale (S Frankl, F Shiere, H Fogels, 1962). Of note, the presence or not of the same dental provider did not influence cooperation’s levels. At one-year follow-up the compliance was preserved, excepting for one child.

#### 4.2.4 AAC and general medical examinations

Addressing challenging behaviors exhibited by children with ASD during medical visits, healthcare providers of University of Iowa Hospitals and Clinics developed visual schedules to support 17 children with ASD in different healthcare examinations (e.g. administration of oxygen, x-ray, EEG, injections, finger sticks) (Chebuhar et al., 2013). Almost 87.5% of the medical staff and 77.8% of caregivers interviewed reported a decrease in children’s anxiety levels and more tolerable visits using AAC boards. Further, parents welcomed the idea of using visual boards, expressing a sense of reassurance.

Thunberg and collaborators (Thunberg, Törnhage, et al., 2016) underlined the potential of AAC in boosting children’s compliance in 7 out of 25 children (age range 3-15; 17 males and 7 females) with communicative disabilities undergoing day surgery. AAC tools were used for preparing children at home and during healthcare procedures. Although anxiety levels, detected with the State-Trait Anxiety Inventory (STAI) (Nilsson et al., 2012), did not statistically change neither among AAC group and controls or their parents, caregivers of children involved in the AAC program with pictograms for preparatory support showed a lower median STAI score on the upon arrival at the hospital (mean score of 9 compared to 11 of parents’ controls).

#### 4.2.5 Perspectives on AAC

Thunberg and colleagues (Thunberg, Buchholz, et al., 2016) investigated caregivers’ ideas on AAC interviewing 10 mothers of children with communicative needs. AAC tools were perceived as valuable for supporting personalized care, safety, and direct interactions; moreover, they perceived as facilitating children’s expression of their needs, understand procedures, and communicate more effectively with healthcare providers. AAC was noted to decrease children’s distress during hospital stays and enhance communication effectiveness in medical settings. Furthermore, a qualitative search based on interviews of 20 Swedish nurses underlined health providers’ awareness of AAC in facilitating the care of children with ID. Specifically, nurses claimed the importance of AAC tools in assessment and understanding care needs. However, interviews highlighted the lack of professional education in implementing AAC in medical routine (Appelgren et al., 2022).

## 5 AAC protocol development for brain stimulation

Since to the best of our knowledge no one has developed AAC protocol yet for sustaining brain stimulation procedures, the creation of AAC aids represents a unique opportunity to better explicate research stages to children with NDDs. Although brain stimulation is an evidence-based research practice increasingly known among clinicians and researchers, it would be possible that brain stimulation procedures and tools may not be familiar to children with NNDs and their families, thereby necessitating specific support, such us visual aids, for implementing clinical procedures.

### 5.1 AAC aids

Pictures describing different clinical and research procedures have been specifically developed by a team made of four psychologists and a speech pathologist and assembled into visual schedules. Features of realism, iconicity (association between a symbol and its referent), ambiguity, complexity, figure-ground differential, perceptual distinctness, acceptability, efficiency, color and size discriminate pictograms were followed (Lloyd et al., 1998; Martínez-Santiago et al., 2018; Schlosser, 2003). Specific attention was placed on guessability or transparency to conversational partners and ease of acquisition by combining pictures to traditional orthography (e.g., printed words) (Martínez-Santiago et al., 2018). Visual aids were specifically created thanks to a graphic that designed and drew colored pictograms with the help of a speech pathologist. Two psychologists and a speech pathologist checked the words supporting images (Figure 2).

**Figure 2.**
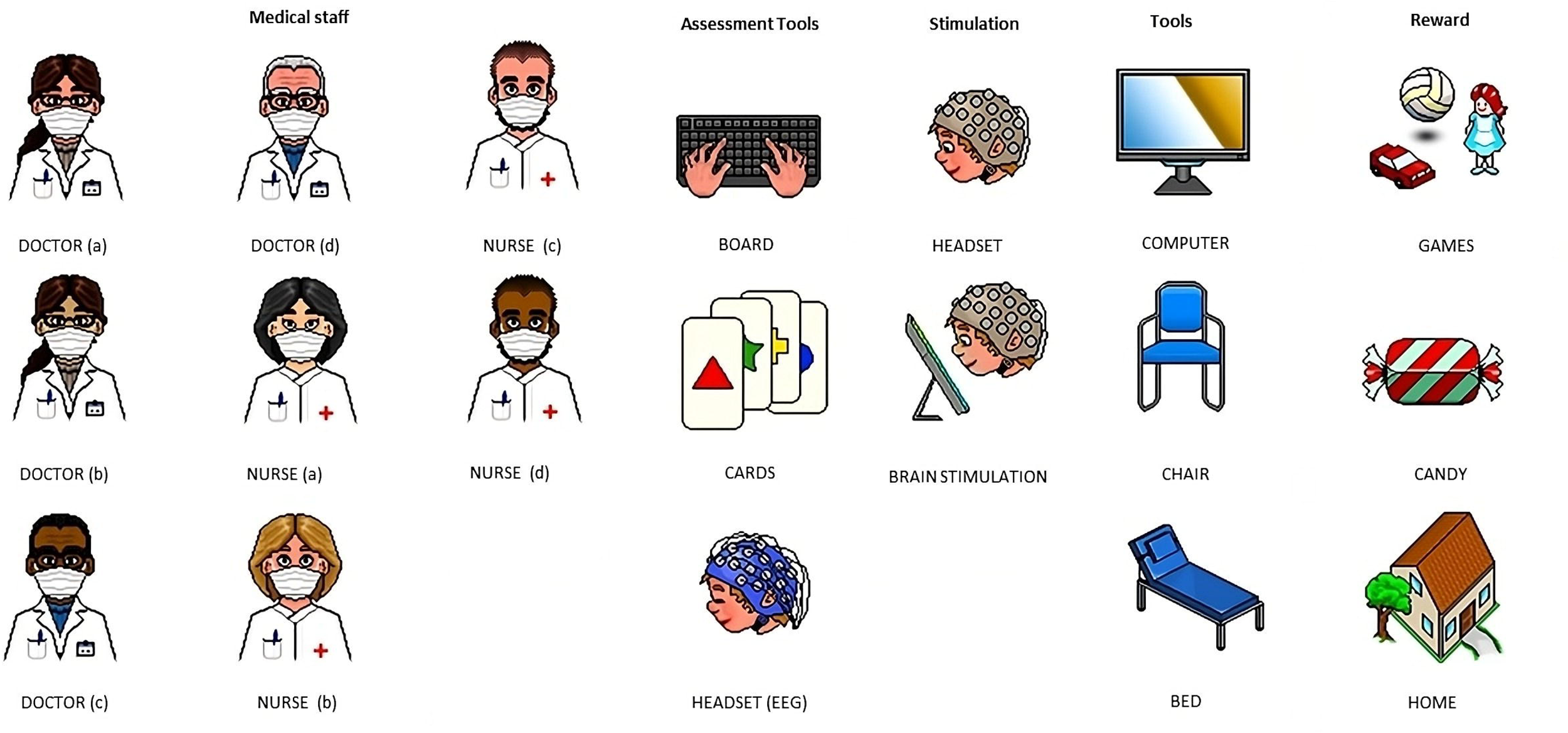
AAC aids for describing research procedures. The researcher selects and assembles the pictograms based on the procedure to be implemented. The researcher assembles each pictogram to create a visual board describing the peculiar research procedure involving the patient.

Pictograms (3×3 cm) have been arranged in a horizontal format with printed words below them. To depict day hospital activities, specifically involving brain stimulation, a sequence of four to six pictures has been attached from left to right. Each picture has Velcro attached to its back, facilitating the creation of sequences (Figure 3).

**Figure 3.**
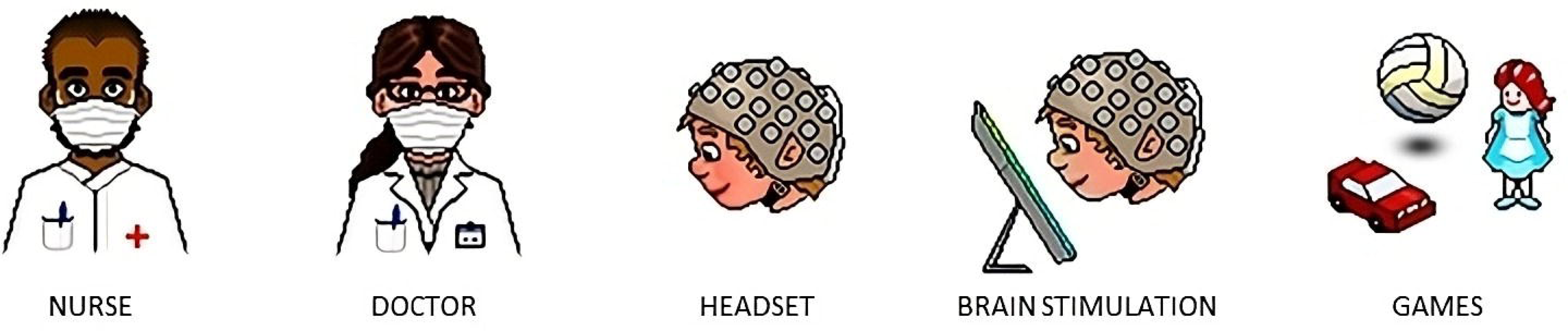
Example of AAC table for brain stimulation procedures. The researcher should touch each visual aid and pronounce the corresponding word to illustrate the procedure. The current AAC aid describe each step to perform a session of brain stimulation, as follows: triage nurse; meeting with the medical staff; putting on the headset; brain stimulation; reward.

Furthermore, for an overall inclusive research practice, we created additional visual aids for monitoring potential sensations linked to brain stimulation (e.g. headache, itch, neck pain). Specifically, pictograms for translating the Safety Questionnaire (Brunoni et al., 2011) into AAC were created for collecting reliable information from children with NDDs after undergoing brain stimulation. Visual aids depicting possible effects linked to brain stimulation and a chart to express sensations’ gradient were developed (Figure 4).

**Figure 4.**
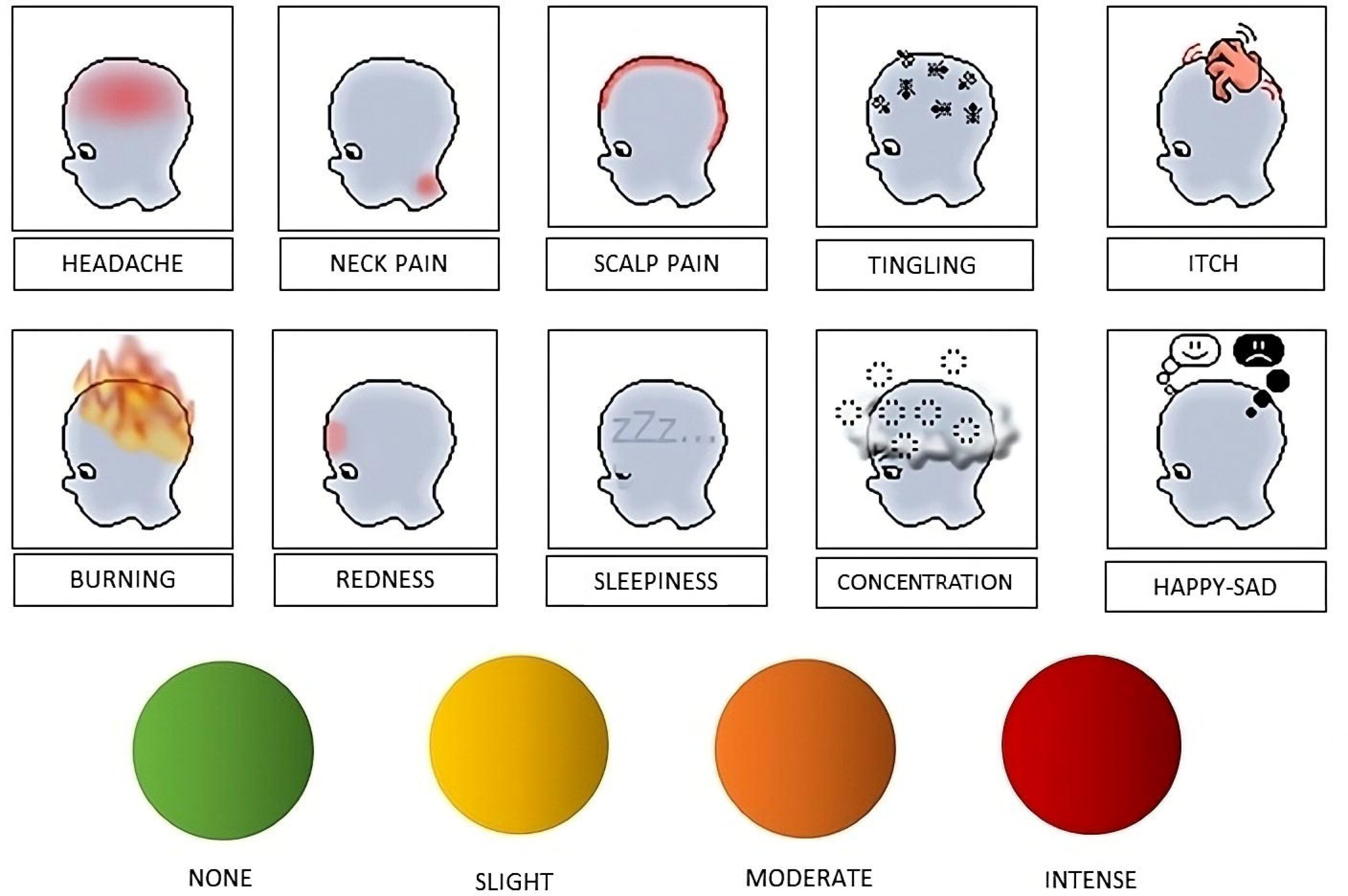
AAC version of the Adverse Effects Questionnaire by Brunoni et al. (2011). The visual aids reproduce the items from the Safety Questionnaire administered after each session to monitor its possible effects. The researcher should touch each visual aid and pronounce the corresponding word to state potential effects linked to brain stimulation. The patient expresses the level of experienced effects by using colored circles.

## 6 Discussion

Children with NDDs frequently display medical comorbidities requiring regular checkups, which can be challenging to perform due to their communication difficulties. Receptive language anomalies may cause problems in understanding healthcare instructions, leading to frustration, perceived loss of control and consequentially challenging and non-compliant behaviors. Further, expressive language issues may complicate describing needs and seeking assistance, exacerbating noncompliance and negative healthcare experiences. These issues may extend to clinical trials, including brain stimulation, since they often encompass research procedures highly unfamiliar to children with NDDs and their families. AAC may be a valuable resource to overcome these issues, given its potential in supporting individuals in expressing themselves, asking for help and explanations, and conveying their sensations and emotions (Beukelman & Light, 2020). AAC promotes an inclusive perspective, recognizing all individuals as having their own agency and thoughts, regardless of the spectrum of disability and language anomalies.

Therefore, the first aim of the current work was to explore the employment of AAC to support brain stimulation procedures. Our literature review failed to find studies focusing on the use of AAC to foster compliance with research procedures based on brain stimulation procedures and involving children with NDDs.

Clinical scientific research aims to strength and expand knowledge to ensure better well-being for all individuals, even in the presence of NDDs. New technologies are enhancing research strategies with a great potential of transforming health-care delivery and acting on atypical developmental trajectories (Boivin et al., 2015). Hence, reflecting on research procedures is crucial for advancing scientific progress without burdening feasibility and time constraints. Researchers, recognizing the uniqueness and individuality of all people involved in their protocols, should embrace an inclusive and individualized approach combining the methodologic scientific rigor, based on replication on larger and more representative samples, with a clinical-human-centered perspective. As research on NDDs has been undergoing significant transformations in recent years, especially in the areas of early identification and lifelong non-invasive interventions (Olivier et al., 2023), considering the unique needs and priorities of children with NDDs before and during the research process is crucial. Further, the acceptability of new research procedures should be thoroughly explored from the points of view of people involved, including children with NDDs, caregivers, and healthcare workers. Future research would benefit from embracing a more inclusive methodology that encompasses the entire range of symptom severity within NDDs.

Given the lack of studies on the use of AAC to sustain brain stimulation procedures in children with NDDs, we conducted a second literature review to explore the potential of AAC in fostering compliance to medical procedures in children with NDDs.

Most of the included studies (67%) focused on dental care, aiming to encourage pediatric dentists to use AAC to alleviate distress, anxiety, and sustain compliance. Reviewed studies underlined the crucial role of AAC in facilitating efficient communication between children and healthcare providers, improving children’s understanding of medical procedures, reducing distress and anxiety, and associated challenging behaviors, thereby enhancing medical compliance. AAC enables the clear explanation of medical steps, whether invasive or not, by educating, preparing, and anticipating procedures in a simple and intuitive manner. In addition, the use of visual aids can enhance the healthcare experience also for parents by providing reassurance and supporting the explanation of health-related procedures, helping caregivers mitigate and manage challenging behaviors. So far, AAC can restructure the care experience to be less difficult and stressful, reducing the need for parents to use specific containment resources to assist healthcare providers in delivering care. Not only caregivers, but also healthcare providers reported the utility of AAC in improving children’s behaviors and compliance (Appelgren et al., 2022; Thunberg, Törnhage, et al., 2016). However, health workers underlined the importance of undergoing proper AAC trainings to not feel unprepared when using visual tools (Handberg & Voss, 2018). Several limitations of the included studies should be acknowledged. First, none provided information on randomization processes or a priori sample size calculation. Second, most studies had small sample sizes. Importantly, all studies, even those conducted in the same clinical context, used different AAC aids for the same procedures, limiting replicability. Additionally, most research considered outcomes from a qualitative perspective, whereas quantitative methods could be more informative. These methodological issues hinder the generalizability of the results. Further studies with larger samples of individuals with various NDDs, and training for both healthcare workers and caregivers on AAC, are crucial to maximize its benefits. Additional research is needed to advance AAC knowledge and refine its application for individuals with communication needs, particularly in healthcare and research contexts (Light & Mcnaughton, 2015). Finally, it should be noticed that none of the included studies specifically and explicitly focused on pain assessment. Given the difficulty of obtaining verbal information from children with NDDs and communication anomalies, AAC for pain monitoring could significantly enhance their health-related experiences and ensure they receive appropriate and effective pain treatment. However, despite the recognized usefulness of AAC in fostering compliance of children with NDDs with clinical procedures, the creation of AAC tools for depicting pain remains underexplored, and attention should be given to individualizing pain-related vocabulary and illustrative pictograms (Johnson et al., 2016).

To sum, our literature review emphasizes that AAC can help overcome communication barriers for children with NDDs, fostering a positive environment that benefits both children and healthcare professionals by supporting interactions and facilitating efficient intervention procedures. All these positive features can also be extended to brain stimulation procedures, making AAC a particularly suitable tool to support clinical procedures in this field as well. A valuable option for inclusive research practice stays in using AAC with brain stimulation to achieve a patient-centered approach in reality. This practice would enable children with NNDs to more easily express themselves and comprehend their experiences by considering the full spectrum of their verbal abilities.

The creation of shared protocols in both clinical and research practices may promote a systematic use of AAC, enabling a more rigorous analysis of its benefits. As stated by the Center for Disease Control and Prevention (Centers for Disease Control and Prevention, 2024), people with disabilities commonly encounter barriers due to reluctant attitudes from the surrounding environment towards their specific needs. Emphasizing the utility of programs using AAC, the CDC aims to truly embrace and implement social policies’ principles of sustainability and inclusion in delivering effective public health programs.

## 7 Conclusion

Although the use of AAC to sustain brain stimulation procedures has not yet been explored, the present study underscores the advantages of pictures schedule to enhance compliance of children with NDDs undergoing medical examinations. The current review promotes the basis for developing future AAC protocols to improve the quality of healthcare and the equity in clinical and research settings. Moreover, the AAC protocol presented in this paper would contribute to fill the gap on using visual aids across researchers and clinical practitioners.

Addressing compliance with research procedures is essential for delivering high-quality medical care, as it is grounded in research itself. Of note, implementing AAC protocols may be crucial to fully respect ethical principles while advancing scientific knowledge.

## Acknowledgments

We thank Mr Marco Nasini for his valuable contribution in developing AAC pictures.

## Author contributions

Conceptualization, S.P., E.F., and F.Q.; methodology, S.P., F.Q and G.L.; formal analysis, S.P. and G.L.; investigation, S.P., L.C., A.B., S.G. and F.C.; data curation, S.P., E.F., F.C. and G.V.; writing—original draft preparation, S.P., E.F., F.Q., G.L., F.C., L.C., G.V., A.B., S.G., D.M., S.P. and S.V; writing—review and editing, S.P., E.F., F.Q., G.L., F.C., A.B., S.G., L.C. and S.P.; supervision, E.F., D.M., and S.V.; project administration, E.F. and S.V. All authors have read and agreed to the published version of the manuscript.

Statements and declarations Ethical considerations

There are no human participants in this article and informed consent is not required.

## Data availability statement

Data sharing not applicable to this article as no datasets were generated or analyzed during the current study.

